# At-home self-testing of teachers with a SARS-CoV-2 rapid antigen test to reduce potential transmissions in schools: Results of the SAFE School Hesse Study

**DOI:** 10.1101/2020.12.04.20243410

**Authors:** Sebastian Hoehl, Barbara Schenk, Olga Rudych, Stephan Göttig, Ivo Foppa, Niko Kohmer, Onur Karaca, Tuna Toptan, Sandra Ciesek

## Abstract

**Background:** Rapid antigen tests for SARS-CoV-2 became available recently, offering an opportunity to vastly increase testing capacities. Antigen tests offer lower sensitivity than the gold standard, RT-PCR, but rapid sample-to-answer time. High-frequency testing with an antigen test may offset the lower sensitivity, and testing can be done with at-home collection of samples, offering potential benefit in screening efforts. In this study, we set out to evaluate the practical application of self-performed high-frequency antigen test in a school setting.

**Method:** A total of 711 teachers from 86 schools were enrolled in a seven-week study. After instruction, participants tested themselves every 48 hours at home with a rapid antigen test for SARS-CoV-2 (target: nucleocapsid protein) in a self-collected anterior nasal swab. Positive results in the antigen test were confirmed via RT-PCR from the same sample that had been determined to be positive by the study participant. A questionnaire was given to all participants to evaluate whether the test failed to detect infection.

**Findings:** 10 836 tests from 602 teachers were recorded and analyzed. A total of five confirmed cases of viral shedding of SARS-CoV-2 was detected by use of the antigen test. One study participant with a SARS-CoV-2 infection was presymptomatic and four were mildly symptomatic at the time of the antigen test. Sixteen false positive antigen tests (0.15% of all tests) were reported, predominantly when the local incidence in the general population was low. In four cases, the study participant reported that a PCR had detected a SARS-CoV-2 infection, but the antigen test was negative, indicating a false negative result.

**Interpretation:** High-frequency, self-performed rapid antigen tests can detect individuals with a SARS-CoV-2 infection, and therefore potentially reduce transmissions. Testing may be most beneficial when applied during high local incidence of SARS-CoV-2 infections and when mild or atypical symptoms are present. To avoid a high rate of false positive results, a test with optimized specificity should be used.

**Funding:** The study was commissioned and funded by the Hessian Ministry of Education and the Hessian Ministry of Integration and Social Affairs.

## Introduction

In the ongoing pandemic of SARS-CoV-2, test strategies continue to rely mostly on real time (RT-) PCR, a highly sensitive and specific method to detect SARS-CoV-2 RNA. Rapid antigen tests (RATs) are also becoming increasingly available and offer point-of-care testing, with results available within minutes. These tests, that are much more affordable and offer the potential of increasing testing capacities as well as reducing sample-to-answer-time, are limited by a lower sensitivity and specificity when compared to PCR.^1-3^ However, the sensitivity of RATs may be sufficient to detect most infectious cases^2^, especially when testing is performed in high frequency.^4,5^ RATs are usually very simple to perform, and can be combined with self-sampling, to offer at-home testing for non-medical professionals. When using a RAT to test for SARS-CoV-2, anterior nasal swabs have been shown to be a viable alternative to oropharyngeal or throat swabs, when collected by a medical professional,^1^ as well as when the sample was self-collected by a non-medical professional.^3,6-8^

High-frequency testing for SARS-CoV-2 with an RAT is proposed to make some critical environments safer during the pandemic when other non-pharmaceutical interventions are not sufficient to prevent transmissions. Theoretical models have proposed that high-frequency antigen testing may be efficacious to reduce transmissions in such environments.^5,9^ Keeping schools open during the pandemic should be considered a high priority due to potentially detrimental effects of withholding education from children of various age groups. Here, the application of rapid antigen tests could prove to be beneficial.

There is limited experience from real-life application of RATs, especially when testing is performed by a non-medical professional without supervision. The primary aim of this study was to determine whether at-home testing results in true-positive detection of unknown viral shedding.

## Methods

The *SAFE School Hesse Study* (SAFE acronym: German: **SA**RS-CoV-2 **F**rüh**E**rkennung, English: Early detection of SARS-CoV-2) set out to evaluate the practical application of self-testing for SARS-CoV-2 with a RAT, performed by teachers. The primary goal of the study was to evaluate the potential of an RAT to detect SARS-CoV-2 infected individuals and thus to enable necessary infection control measures.

Teachers from primary and secondary schools in three distinct school districts were invited by the Hessian Ministry of Education to participate in the study. The study period was seven weeks, of which three were before and four were after a two-week autumn vacation in Hesse (September 14^th^ to October 4th^nd^ and October 17^th^ to November 15^th^, 2020).

Study participants received written instructions, as well as an instructional video on how to collect an anterior nasal swab, and step-by-step instruction to perform and read out an RAT (RIDA^®^ QUICK SARS-CoV-2 Antigen test, R-Biopharm, Darmstadt, Germany) at home. In the testing process, two reagents were combined with drop bottles, the nasal swabs were irrigated in medium, and a predetermined amount of this sample was transferred to the testing tube with the reagents using a single-use pipet. After 10 minutes of incubation, a lateral flow test strip was added to the test tube. The visual result readout was performed after an additional 10 minutes. Study participants were instructed to observe whether the control band was visible to determine the validity of the result, and to determine whether the test band was visible to determine a positive (test band visible) or negative (test band not visible) result. Study participants were instructed to record all tests on a standardized form.

Study participants were provided with the telephone number to the study hotline as well as a designated email address in case any support was needed. In case of positive or inconclusive test readouts, participants were asked to call the study hotline for further instructions. When a sample was deemed or suspected to be positive by the study participant, the sample medium from which material was inserted into the antigen test by the study participant was collected. In the laboratory (Institute for Medical Virology, Goethe University Frankfurt), a SARS-CoV-2 RT-PCR was performed, using either Roche Cobas^®^ SARS-CoV-2 (Roche diagnostics, Basel, Switzerland), Alinity m^®^ SARS-CoV-2 Assay (Abbott Laboratories, Abbott Park, IL, USA) or Xpert^®^ Xpress SARS-CoV-2 (Cepheid, Sunnyvale, CA, USA). The study participants were asked to self-quarantine until PCR results were conveyed.

At the end of the study, all participants received a questionnaire to determine whether they encountered any difficulty in the testing procedure, and to record whether they were diagnosed with a SARS-CoV-2 infections but were tested negative with the RAT.

When a study participant was quarantined due to exposure to SARS-CoV-2, positive test results were not evaluated. When a positive antigen test was recorded, either true or false, no further RAT tests were analyzed from the respective study participant.

### Funding and ethical approval

This study was commissioned and funded by the Hessian Ministry of Education and the Hessian Ministry for Integration and Social Affairs. The study protocol was approved by the ethics board of the University Hospital, Goethe University, Frankfurt am Main, Germany (No. 20-899).

## Results

### Characterization of study participants

711 teachers participated in the study. A total of 602 teachers (86.7%) from 85 schools provided records from the RATs. The age range was 21 to 67 years. A total of 10836 tests were recorded (mean: 18 tests per study participant). 10 768 of these tests (99.37%) were recorded to have been valid and negative, 47 (0.43%) were recorded as invalid and 21 (0.19%) as positive (either true or false).

### True positive results

A true positive antigen test result was confirmed by RT-PCR in five teachers during the study period (table 1). A prior or ongoing infection with SARS-CoV-2 was not known in any of these participants at the time of the antigen test. All five true positive cases occurred when the local 7-day incidence in the school district was higher than 100 cases / 100 000 inhabitants (mean 7-day incidence at the time of true positive test result: 252.66 cases / 100 000 inhabitants). At the time of testing, four of them were symptomatic, and one was presymptomatic. No asymptomatic infection with SARS-CoV-2 was detected in this study. The Cycle-threshold (Ct) values for the five RAT-positive samples ranged 17.6 and 20.2.

### False positive results

In 16 cases, a positive RAT result was determined by the study participant but could not be confirmed by RT-PCR. 13 of these tests were conducted when the 7-day local incidence was below 40 cases / 100 000 inhabitants, and three occurred with a 7-day local incidence between 171 and 213 cases / 100 000 inhabitants. 12 of the 16 false positive tests occurred while the study participant was asymptomatic, and four while symptoms were reported by the study participant (table 1). In 10 of these 16 cases, the false positive result occurred in the very first test the teacher performed.

Protein A of S*taphylococcus aureus* and other coagulase positive Staphylococcus species are known to bind the Fc region of IgG and which can lead to false positive reactions in lateral flow test systems.^10^ Therefore, seven false positive anterior nasal swab samples were analyzed for the presence of S. *aureus* by culture and PCR (target sequence: nuc and MREJ). Six out of seven samples were positive by PCR and four out of seven samples were culture-positive.

### False negative results

For four teachers, a false negative result in the antigen test was assumed, as they reported to have received a positive test result by SARS-CoV-2 PCR from a swab that had been collected by a medical professional during the time of high-frequency self-testing with the RAT. Three of these events occurred during high incidence of SARS-CoV-2 (171, 213 and 253 cases / 100 000 inhabitants, respectively). Two of these study participants were symptomatic and one was presymptomatic at the time of the positive PCR test result (table 1). For one case it was not reported when the false negative test occurred, and if symptoms were present. In one case, the RAT later was positive seven days after the RT-PCR result.

### Local incidence during the study period

The 7-day local incidence of SARS-CoV-2 infections in the general population was 9 to 348 cases per 100 000 inhabitants.

## Discussion

We evaluated the practical application of at-home, high-frequency RATs for SARS-CoV-2 by teachers to prevent SARS-CoV-2 transmission in schools.

The test used in this study is a lateral flow SARS-CoV-2 rapid antigen test. While the overall sensitivity of this test was only 50.0 to 77.6% in a recent study, it was 88.2% when samples were used with Ct values below 28, a range that is typically seen during the first week of a SARS-CoV-2 infection, when the patient is likely infectious. Cumulative specificity for the test has been reported to be 94.85%.^11^

The objective of the study was to determine whether at-home testing results in true-positive detection of unknown viral shedding, causing the potentially infectious teacher to become aware of an infection with SARS-CoV-2, and therefore preventing potential transmissions in school. This outcome was archived in five out of 602 study participants. These individuals had a high viral load in the self-collected nasal swab (Ct values 17.6 to 20.2), indicating they were likely infectious. Possible onward transmissions in the school or in a private setting may have been prevented. All but one of these participants were mildly symptomatic at the time of testing, but only one reported fever at the time of testing. This demonstrates that RAT testing is especially useful when symptoms are present, even in the absence of fever. In one case, headache and fatigue were the only reported symptoms, which highlights the need to test for SARS-CoV-2 even in cases with symptoms that are very prevalent in the general population that may not be immediately attributed to COVID-19.

A total of 16 false positive results were recorded by the study participants. Notably, 13 of the 16 false positive tests occurred while the local 7-day incidence of SARS-CoV-2 infections was below 100 cases / 100 000 inhabitants, demonstrating the implications of a low positive predictive value of the RAT during low infection prevalence. To avoid the occurrence of false positive results, large-scale testing could be applied during periods of high incidence, and a test with higher specificity should be used in such screening efforts.

In this study, the test material was a nasal swab, which is demonstrated to offer high diagnostic sensitivity, which could also be demonstrated for self-collected samples by non-medical professionals.^3,6,7^ We observed 16 false positive results; however, when compared to the total number of study participants, this occurred only in 2.66% of all study participants. As anticipated, the rate of false positive results decreased with a rising prevalence of SARS-CoV-2 that occurred throughout the study period (table 1). The decrease in the false positivity rate may also indicate that false positive results occurred due to a permanent host factor that resulted in these study participants to drop out of the study as it progressed. One such conceivable factor is the nasal colonization with *Staphylococcus aureus*, which could result in false positive result due to interference with the lateral flow assay through the binding of immunoglobulin by staphylococcal protein A (SpA).^12^ Indeed, in six out of seven false positive samples, the presence of S*taphylococcus aureus* could be confirmed.

When reviewing the records, very few tests yielded an invalid result (47 out of 10 836 tests, 0.43%), indicating that the RAT could be performed satisfactory by the study participants, who could call a hotline in case any technical or medical support was needed. It is imperative that, when the test is performed without supervision of a medical professional, support is easily accessable.^13^

Presumably, four out of 10 836 tests were false negative, which might be due to insufficient swabbing technique, an error in testing, result readout, or low viral load in anterior nasal sampling.

This study has several limitations. All participants were educators in primary or secondary schools, and therefore had a higher education. Not all teachers at the schools that participated in the study performed the test every 48 hours throughout the entire study period, and not all teachers from the schools participated in the study. Furthermore, no students from these schools were tested, and transmissions in the participating schools were not examined. As no students were enrolled in the study, we also do now know whether the approach of high-frequency, self-performed testing is beneficial in children.

In conclusion, we could demonstrate that a high frequency, self-performed antigen tests by teachers can result in the detection of individuals who are probably infectious, and therefore could contribute to making the school environment safer during the pandemic.

Testing for SARS-CoV-2 is especially important when symptoms are present, which may also be mild or atypical. As infections may also be missed by use of an antigen test, all non-pharmaceutical interventions should be applied regardless of the use of rapid antigen tests.

The authors received positive feedback from the study participants throughout the study, a large majority of whom experienced the study participation with regular testing for SARS-CoV-2 to be reassuring when working in schools during the pandemic.

## Data Availability

All relevant data are available upon reasonable request.

## Appendix

**Table 1:** Characterization of all true positive, false positive and false negative antigen tests results determined in the SAFE School Study. Ag.: Antigen, n.a.: Not applicable. * result interpretation by the study participant at home. + positive: test band weaker than control band. ++ positive: test band as strong as control band. +++ positive: test band stronger than control band. ^1)^ Cepheid Xpert Xpress^®^ SARS-CoV-2 ^2)^ Abbott Alinity M^®^ SARS-CoV-2 ^3)^ Roche Cobas^®^ SARS-CoV-2 on the Cobas^®^ 6800 ^4)^ PCR done elsewhere # ORF-region

| No. | Study week | Result of Ag Test at home* | Result SARS-CoV-2 PCR | Ct value | Classification of Ag Test Result | Local 7-day-incidence (school district) | Symptoms of COVID-19 |  | Symptom Classification at time of Ag test |
| --- | --- | --- | --- | --- | --- | --- | --- | --- | --- |
|  |  |  |  |  |  |  | Symptoms at time of Ag test | Days of symptoms prior to Ag test |  |
| 1 | 1 | + | negative<sup>1)</sup> | n.a. | false positive | 17.71 | none | n.a. | asymptomatic |
| 2 | 1 | + | negative<sup>2)</sup> | n.a. | false positive | 17.71 | sore throat | 0 | symptomatic |
| 3 | 1 | ++ | negative<sup>3)</sup> | n.a. | false positive | 17.71 | none | n.a. | asymptomatic |
| 4 | 1 | ++ | negative<sup>3)</sup> | n.a. | false positive | 14.58 | none | n.a. | asymptomatic |
| 5 | 1 | + | negative<sup>1)</sup> | n.a. | false positive | 17.71 | none | n.a. | asymptomatic |
| 6 | 2 | + | negative<sup>3)</sup> | n.a. | false positive | 11.75 | none | n.a. | asymptomatic |
| 7 | 2 | +++ | negative<sup>3)</sup> | n.a. | false positive | 24.45 | none | n.a. | asymptomatic |
| 8 | 2 | ++ | negative<sup>3)</sup> | n.a. | false positive | 24.45 | none | n.a. | asymptomatic |
| 9 | 2 | +++ | negative<sup>1)</sup> | n.a. | false positive | 31.47 | none | n.a. | asymptomatic |
| 10 | 3 | +++ | negative<sup>3)</sup> | n.a. | false positive | 28.10 | none | n.a. | asymptomatic |
| 11 | 3 | + | negative<sup>3)</sup> | n.a. | false positive | 39.91 | rhinitis | 4 | symptomatic |
| 12 | 3 | + | negative<sup>2)</sup> | n.a. | false positive | 28.10 | none | n.a. | asymptomatic |
| 13 | 3 | ++ | negative<sup>3)</sup> | n.a. | false positive | 39.91 | none | n.a. | asymptomatic |
| 14 | 4 | +++ | positive<sup>3)</sup> | 18.9<sup>#</sup> | true positive | 108.76 | fever, cough, head ache, body ache | 3 | symptomatic |
| 15 | 5 | +++ | positive<sup>2)</sup> | 20 | true positive | 267.12 | headache, fatigue | 2 | symptomatic |
| 16 | 5 | + | negative<sup>2)</sup> | n.a. | false positive | 211.35 | sore throat | 0 | symptomatic |
| 17 | 6 | ++ | negative<sup>3)</sup> | n.a. | false positive | 213.60 | abdominal pain | 3 | symptomatic |
| 18 | 6 | - | positive<sup>4)</sup> | external test | false negative | 213.60 | none | 0 | presymptomatic |
| 19 | 6 | +++ | positive<sup>3)</sup> | 17.6<sup>#</sup> | true positive | 275.56 | fatigue, body ache | 0 | symptomatic |
| 20 | 6 | + | positive<sup>3)</sup> | 19.7<sup>#</sup> | true positive | 139.45 | cough, nasal congestion, loss of smell and taste, headache | 2 | symptomatic |
| 21 | 7 | ++ | positive<sup>3)</sup> | 20.2<sup>#</sup> | true positive | 252.66 | none | n.a. | presymptomatic |
| 22 | 7 | negative | positive<sup>4)</sup> | external test | false negative | 252.66 | congested nose | 2 | symptomatic |
| 23 | 7 | + | negative<sup>3)</sup> | n.a. | false positive | 171.51 | none | n.a. | asymptomatic |
| 24 | 7 | negative | positive<sup>4)</sup> | external test | false negative | 171.51 | nausea | 2 | symptomatic |

## Acknowledgement

We would like to thank Marhild Kortenbusch, Regine Jeck, Jessica Gille and Lena Marie Pompe for technical support and RT-PCR testing.

## Literature

1. Abdulrahman A, Mustafa F, AlAwadhi AI, Alansari Q, AlAlawi B, AlQahtani M. Comparison of SARS-COV-2 nasal antigen test to nasopharyngeal RT-PCR in mildly symptomatic patients. medRxiv 2020: 2020.11.10.20228973.

2. Corman VM, Haage VC, Bleicker T, et al. Comparison of seven commercial SARS-CoV-2 rapid Point-of-Care Antigen tests. medRxiv 2020: 2020.11.12.20230292.

3. Lindner AK, Nikolai O, Kausch F, et al. Head-to-head comparison of SARS-CoV-2 antigen-detecting rapid test with self-collected anterior nasal swab versus professional-collected nasopharyngeal swab. medRxiv 2020: 2020.10.26.20219600.

4. Mina MJ, Parker R, Larremore DB. Rethinking Covid-19 Test Sensitivity — A Strategy for Containment. New England Journal of Medicine 2020.

5. Larremore DB, Wilder B, Lester E, et al. Test sensitivity is secondary to frequency and turnaround time for COVID-19 screening. Sci Adv 2020.

6. Tu YP, Jennings R, Hart B, et al. Swabs Collected by Patients or Health Care Workers for SARS-CoV-2 Testing. N Engl J Med 2020; 383(5): 494–6.

7. Altamirano J, Govindarajan P, Blomkalns AL, et al. Assessment of Sensitivity and Specificity of Patient-Collected Lower Nasal Specimens for Sudden Acute Respiratory Syndrome Coronavirus 2 Testing. JAMA Netw Open 2020; 3(6): e2012005.

8. Lee RA, Herigon JC, Benedetti A, Pollock NR, Denkinger CM. Performance of Saliva, Oropharyngeal Swabs, and Nasal Swabs for SARS-CoV-2 Molecular Detection: A Systematic Review and Meta-analysis. medRxiv 2020: 2020.11.12.20230748.

9. Paltiel AD, Zheng A, Walensky RP. Assessment of SARS-CoV-2 Screening Strategies to Permit the Safe Reopening of College Campuses in the United States. JAMA Network Open 2020; 3(7): e2016818.

10. Cronin UP, Girardeaux L, O’Meara E, Wilkinson MG. Protein A-Mediated Binding of Staphylococcus spp. to Antibodies in Flow Cytometric Assays and Reduction of This Binding by Using Fc Receptor Blocking Reagent. Applied and Environmental Microbiology 2020; 86(17).

11. Corman VM, Toptan T, et al. Evaluation of a SARS-CoV-2 rapid antigen test: potential to help reduce community spread? Journal of Clinical Virology 2020. accepted for publication

12. Ljungberg UK, Jansson B, Niss U, Nilsson R, Sandberg BEB, Nilsson B. The interaction between different domains of staphylococcal protein a and human polyclonal IgG, IgA, IgM and F(ab’)2: Separation of affinity from specificity. Molecular Immunology 1993; 30(14): 1279–85.

13. Rafiei Y, Mello MM. The Missing Piece — SARS-CoV-2 Testing and School Reopening. New England Journal of Medicine 2020.

